# Visceral adipose macrophage content does not associate with body mass index or systemic inflammation in COVID-19: an autopsy study

**DOI:** 10.1101/2023.08.21.23294361

**Authors:** Steven H. Su, Yael R. Nobel, Sepideh Besharati, Armando del Portillo, Michaela R. Anderson, Daniel E. Freedberg

## Abstract

**Introduction:** Adiposity, especially visceral adiposity with elevated body mass index (BMI), is associated with a hyperinflammatory syndrome and poor outcomes in patients with COVID-19. In other diseases such as obesity, type 2 diabetes, and rheumatoid arthritis, systemic inflammation is driven directly by visceral adipose macrophages which release pro-inflammatory cytokines. Currently it is unknown whether visceral adipose tissue macrophage content may similarly explain the observation that COVID-19 patients with elevated BMI are at risk for a hyperinflammatory syndrome and death.

**Methods:** This was a retrospective study of hospitalized adults who died of COVID-19 between March 2020 and June 2020 and underwent autopsy. Visceral adipose tissue macrophage content was quantified by histological staining of visceral adipose tissue samples with CD68, using pericolic fat gathered at autopsy from each subject. Clinical data including inflammatory markers such as erythrocyte sedimentation rate (ESR), C-reactive Protein (CRP), Troponin, D-dimer, Interleukin-6 (IL-6), and ferritin as well as BMI were collected from electronic medical records.

**Results:** A total of 39 subjects were included in this study. There was no association between BMI and visceral adipose tissue macrophage content (Spearman R=0.025, p=0.88). Additionally, there was no association between adipose tissue macrophage content and any of the systemic markers of inflammation measured including ESR, CRP, Troponin, D-dimer, IL-6, and Ferritin (p>0.05 for all markers).

**Conclusion:** Unlike chronic diseases such as obesity, type 2 diabetes, and rheumatoid arthritis, elevated BMI is not associated with increased visceral adipose tissue macrophage content in patients who died of COVID-19. Additionally, among patients who died of COVID-19, visceral adipose tissue macrophage content is not associated with markers of systemic inflammation. These results suggest that the elevations in systemic markers of inflammation—and the hyperinflammatory syndrome often observed during acute COVID-19—does not directly originate from visceral adipose macrophages as it seems to in chronic disease states.

## INTRODUCTION

Since the onset of the COVID-19 pandemic, numerous studies have demonstrated an association between increased body mass index (BMI) and increased visceral adiposity with poor outcomes in patients with COVID-19 including increased likelihood of hospitalization, intensive care unit (ICU) admission, invasive mechanical ventilation, and death ^1–3^. Obesity, especially when associated with increased visceral adiposity, has been shown to increase systemic inflammation in a variety of illnesses including COVID-19 ^1–8^. More specifically, previous work has demonstrated a positive association between visceral adiposity and serum biomarkers of systemic inflammation including C-Reactive Protein (CRP), Erythrocyte Sedimentation Rate (ESR), Interleukin-6 (IL-6), and Ferritin in a variety of diseases including COVID-19 ^9–14^.

In obesity and diseases other than COVID-19 such as type 2 diabetes mellitus and rheumatoid arthritis, systemic inflammation is driven by visceral adipose macrophages through their direct release of inflammatory cytokines including Tissue Necrosis Factor-α (TNF-α) and IL-6 ^13,15–18^. Interestingly there is also an established association between elevations in inflammatory serum biomarkers and the severity of COVID-19 illness ^14^ yet there remains a question of whether adiposity and specifically visceral adipose macrophage content drives systemic inflammation in COVID-19. Given the established relationships between BMI and adipose tissue macrophage content and between adipose tissue macrophage content and systemic inflammation in diseases other than COVID-19, we sought to determine whether elevated BMI is associated with increased visceral adipose macrophage content and whether increased visceral adipose macrophage content is associated with increased serum biomarkers of inflammation in patients with COVID-19.

In this study, we quantified adipose tissue macrophage content in patients who died from COVID-19 through autopsy and also obtained relevant exam and laboratory values including BMI and serum inflammatory markers including ESR, CRP, Troponin, D-dimer, IL-6, and Ferritin to investigate the potential relationships between BMI and adipose macrophage content and between adipose macrophage content and systemic inflammation.

## MATERIALS AND METHODS

### Patient Population

Subjects included in this study tested positive for SARS-CoV-2 by nasopharyngeal PCR and subsequently died of COVID-19 between March 2020 and June 2020 at Columbia University Irving Medical Center. All subjects were age 18 or older and underwent autopsy at the request of next of kin. All protocols utilized in this study were approved by the Columbia University Institutional Review Board and by the Columbia University COVID-19 Biobank (CUB).

### Quantification of visceral adipose tissue macrophage content

Paraffin-embedded tissue sections of colon from 45 COVID-19 autopsy cases were retrieved from the Columbia University Department of Pathology. The microscopic slides from each block were immunolabeled with an antibody against CD68 (Agilent, M087629-2) diluted at 1:50 in antibody diluent (Agilent, S080983-2) per manufacturer’s instructions. Each slide was digitally scanned using a Leica SCN400 whole slide digital imaging system (Leica Biosystems, USA/Germany) at 20X magnification. Using the Aperio ImageScope program (Leica Biosystem Pathology Imaging Inc, Buffalo Grove, IL), images were analyzed. Four areas with pericolic adipose tissue were selected randomly. The number of positive CD68 macrophages were quantified within the selected areas at highest magnification. The median number of CD68-positive macrophages was computed as the visceral adipose macrophage content for each patient.

### BMI, Laboratory Values and Demographics

BMI, date of death, most recent ESR, CRP, Troponin, D-dimer, IL-6 and Ferritin serum laboratory values prior to death and demographics including age, sex, and race were obtained from electronic medical records for each subject. Our rationale for using the most recent laboratory values prior to death was that these values would be more likely to reflect visceral adipose tissue macrophage content compared to (e.g.) peak values for the same labs which may have been from weeks prior. In a secondary analysis, we also examined peak values of the same inflammatory markers during the hospitalization.

### Statistical Analysis

Spearman correlation and p-values were computed for continuous data. For analysis of serum inflammatory markers values (ESR, CRP, Troponin, D-dimer, IL-6 and Ferritin), patients were grouped into tertiles based on visceral adipose macrophage content and p-values were computed using a Kruskal-Wallis test to test for differences within each serum inflammatory marker based on tertiles of macrophage content. All statistical analysis was performed using RStudio software version 2021.09.0-351 running R version 4.1.1.

## RESULTS

### Study Population

Out of 45 patients who underwent autopsy, 6 were excluded because no BMI was recorded during the hospitalization or at autopsy leaving 39 patients for inclusion in the study. Clinical characteristics of these subjects are shown in **Table 1**. The median age was 72 with interquartile range (IQR) 67 - 76. The majority of subjects were male and Hispanic with a median BMI of 26.5 kg/m^2^ (IQR 24.7 – 29.8) and a median adipose macrophage count of 11.5 macrophages/high power field (HPF) (IQR 8.25 - 13.5).

**Table 1.** Demographic and obesity-related characteristics among 39 patients with COVID-19 who died and underwent autopsy.

| **Characteristics** | **Total (n=39)** |
| --- | --- |
| Age, median (IQR) | 72 (67 - 76) |
| Sex, n (%) |  |
| Male | 26 (66.7%) |
| Female | 13 (33.3%) |
| Race/Ethnicity, n (%) |  |
| Hispanic | 25 (64.1%) |
| Black Non-Hispanic | 2 (5.1%) |
| White | 1 (2.6%) |
| Other | 11 (28.2%) |
| BMI, median (IQR) | 26.5 (24.7 - 29.8) |
| Visceral Adipose Tissue Macrophage Content, median (IQR) | 11.5 (8.25 - 13.5) |

IQR (Interquartile Range)

### BMI and adipose tissue macrophage content in patients who died of COVID-19 are not associated

To quantify visceral adipose tissue macrophage content, pericolic visceral adipose tissue was obtained from each subject at autopsy. Tissue sections were stained with both Hematoxylin and Eosin as well as with CD68, a macrophage marker. Representative images of staining are shown in **Figure 1 A-B**. To examine the potential association between BMI and adipose tissue macrophage content, we plotted BMI versus adipose tissue macrophage content (**Figure 1 C**). Upon visual inspection there appeared to be no clearly discernable relationship between the two and this apparent lack of association was supported by a Spearman correlation coefficient near zero with a non-significant p-value (R=0.025, p=0.88), suggesting the absence of a monotonic association between visceral adipose macrophage content and BMI in patients who died of COVID-19.

**Figure 1.**
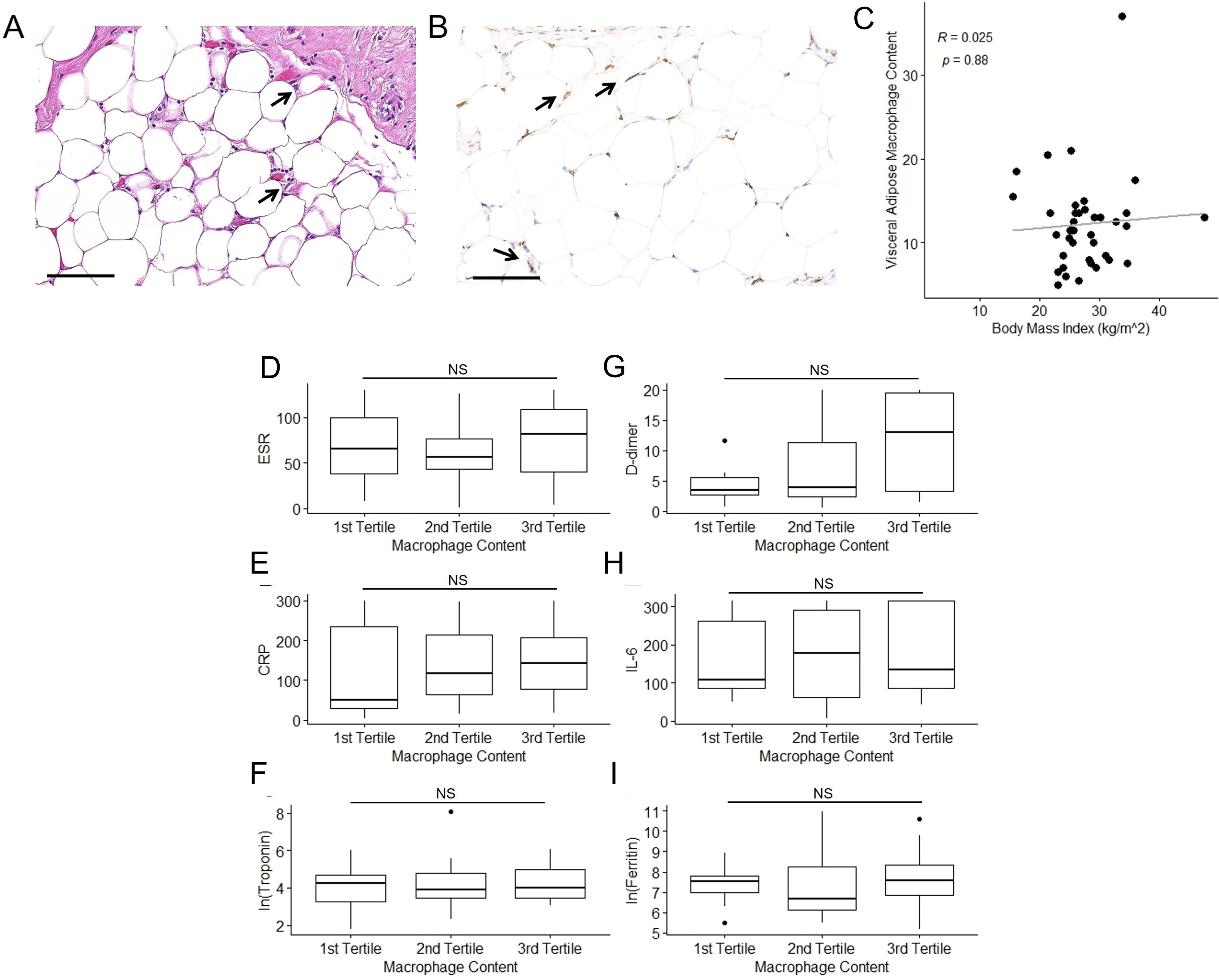
***(A-B)*** Representative images of hematoxylin and eosin ***(A)*** and CD68 ***(B)*** stained visceral adipose tissue sections. Arrows indicate macrophages. Scale bar is 100 μm in ***(A)*** and ***(B)***. ***(C)*** Scatterplot of body mass index plotted versus visceral adipose macrophage content. Line of best fit indicated by grey line. Spearman correlation coefficient (R) and p-value are shown. ***(D-I)*** Laboratory values for Erythrocyte Sedimentation Rate (ESR), C-reactive Protein (CRP), Troponin, D-dimer, Interleukin-6 (IL-6) and Ferritin are stratified by tertiles of visceral adipose tissue macrophage content in ***(D-I)***, respectively. NS indicates non-significant difference as determined via Kruskal-Wallis statistical testing.

### Adipose tissue macrophage content and systemic inflammatory markers are not associated in patients who died of COVID-19

To investigate the potential association between visceral adipose tissue macrophage content and systemic inflammation in COVID-19, the most recent serum ESR, CRP, Troponin, D-dimer, IL-6 and Ferritin values prior to death were obtained for each study subject. In some instances, patients did not have all 6 of these lab values recorded in the electronic medical record and in these cases the remaining available lab values were used. The number of subjects for which each lab value was available is shown in **Table 2** while the median days prior to death for which each of these lab values was measured is shown in **Table 3**. Overall, 58.3% of lab values were within two days of death.

**Table 2.** Serum inflammatory markers, stratified by visceral adipose macrophage content.

| Inflammatory Markers | Visceral Adipose Macrophage Content<br>Median (IQR) |  |  |  | p-value |
| --- | --- | --- | --- | --- | --- |
|  | Overall | 1st Tertile | 2nd Tertile | 3rd Tertile |  |
| ESR (n = 34) | 69<br>(47 - 101.5) | 65<br>(38 - 100) | 56.5<br>(42.7 - 76.8) | 81<br>(40.5 - 109) | 0.69 |
| CRP (n = 35) | 99.7<br>(40.6 - 214) | 51.3<br>(28.4 - 236) | 118<br>(63.1 - 214) | 142<br>(78.7 - 207) | 0.70 |
| Troponin (n = 37) | 63<br>(31 - 112) | 69<br>(27 - 107) | 51.5<br>(31 - 118) | 55<br>(31.5 - 154) | 0.92 |
| D-dimer (n = 33) | 3.9<br>(2.6 - 13) | 3.4<br>(2.7 - 5.5) | 3.9<br>(2.3 - 11.3) | 13<br>(3.2 -19.6) | 0.20 |
| IL-6 (n = 31) | 134.7<br>(85.7 - 315) | 108<br>(87.1 - 261) | 177.3<br>(61.9 – 292) | 135<br>(87 - 315) | 0.89 |
| Ferritin (n = 34) | 7,450<br>(665 - 3,550) | 1,880<br>(1,050 - 2,430) | 806<br>(461 – 4,090) | 1,970<br>(944 – 4,120) | 0.72 |

ESR (Erythrocyte Sedimentation Rate), CRP (C-Reactive Protein), IL-6 (Interleukin-6); p-value computed with Kruskal-Wallis test

**Table 3.**
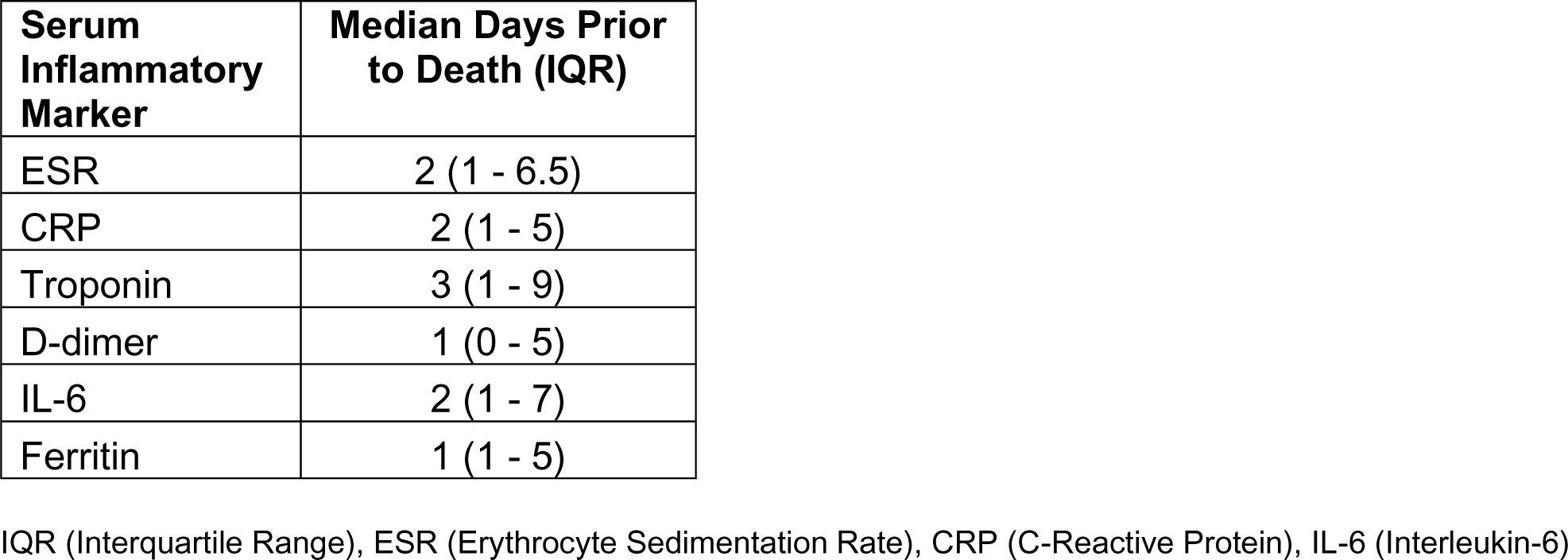
Timing of serum inflammatory markers relative to death.

| **Serum Inflammatory Marker** | **Median Days Prior to Death (IQR)** |
| --- | --- |
| ESR | 2 (1 - 6.5) |
| CRP | 2 (1 - 5) |
| Troponin | 3 (1 - 9) |
| D-dimer | 1 (0 - 5) |
| IL-6 | 2 (1 - 7) |
| Ferritin | 1 (1 - 5) |

IQR (Interquartile Range), ESR (Erythrocyte Sedimentation Rate), CRP (C-Reactive Protein), IL-6 (Interleukin-6)

Subjects were then stratified into tertiles based on their visceral adipose tissue macrophage content. For each marker of systemic inflammation (ESR, CRP, Troponin, D-dimer, IL-6, and Ferritin) the median and IQR were computed within each tertile as well as all subjects together. This data is shown in **Table 2** and **Figure 1 D-I**. For all 6 markers of inflammation, there was no significant differences between tertiles (Kruskal-Wallis p-values >0.05 for all, **Table 2**). Additionally, when comparing visceral adipose macrophage content to each marker of systemic inflammation, there were no correlations (Spearman coefficients all >0.17 with p >0.05). When this analysis was repeated using peak laboratory values obtained at any time during the index hospitalization, the results were substantively unchanged (data not shown).

## Discussion

Adiposity, especially visceral adiposity with increased BMI, has been associated with elevations in systemic inflammation in a variety of diseases including COVID-19 ^4–8^. In diseases other than COVID-19 such as type 2 diabetes mellitus and rheumatoid arthritis, systemic inflammation is in part driven by adipose tissue macrophages through the release of inflammatory cytokines; in these disease states, the degree of inflammation has been shown to be proportional to visceral adipose tissue macrophage content ^13,15–18^. In this study of patients who died of COVID-19, however, we found no association between BMI and visceral adipose macrophage content and also no association between visceral adipose macrophage content and markers of systemic inflammation in. To our knowledge, this is the first study to directly examine adipose tissue macrophage content and systemic inflammation in COVID-19.

Our results are surprising given that adiposity and increased BMI are associated with poor outcomes and increased peak levels of markers of systemic inflammation in COVID-19 ^1–3,7,8,14^. Indeed, one of the striking clinical features of the early pandemic was the hyperinflammatory state that often accompanied severe COVID-19 and led to death, especially in very obese patients. One explanation for the discrepancy between our findings and the previously observed associations between adiposity and BMI with increased systemic inflammation in COVID-19 is that the clinical observations come from living patients whereas our current study is limited to patients at autopsy ^7^. Additionally, our patients were older and had a slightly lower median BMI compared to patients in previous studies that showed strong associations between systemic inflammation and obesity in COVID-19 ^7^. It is possible that obesity (and visceral adipose tissue macrophage content) are crucial to inflammation in young but not old patients with COVID-19.

Another potentially surprising finding is that, unlike in patients with type 2 diabetes, among patients who died of COVID-19, there was no association between BMI and visceral adipose tissue macrophage content. This suggests the possibility that visceral adipose tissue and visceral adipose tissue macrophages may not serve as the primary source of systemic inflammation in severe COVID-19. Speculatively, our results suggest that systemic inflammation may be driven by an element other than visceral adipose tissue macrophages. This idea is supported by the lack of increase in IL-6 in patients with increased adipose macrophage content (**Figure 1 H**, **Table 2**) since IL-6 together with TNF-α have previously been identified as the key inflammatory cytokines released by adipose tissue macrophages in driving systemic inflammation in non-COVID conditions ^4,15–18^. It is possible that the source of systemic inflammation lies outside of adipose tissue, for example in the lungs where the primary pathology of COVID-19 infection occurs. Alternatively, the lack of association between visceral adipose tissue macrophage content and BMI maybe attributable to the relatively low median BMI of our study population.

Our study has several limitations including the inability to study clinical outcomes and lack of standardization in the timing of when laboratory values were collected. Despite this latter limitation, 58% of labs were collected within 48 hours before death. Our cohort had a median age greater than 65 and was predominantly normal weight or overweight (median BMI 26.5, IQR 24.7-29.8 kg/m^2^). Younger and more obese patients may have stronger associations between obesity, visceral adipose tissue macrophage content, and systemic inflammation. Another limitation is that our study population was predominantly male and Hispanic, which may affect generalizability.

In summary, we found no association between BMI and visceral adipose tissue macrophage content among patients who died of COVID-19. Additionally, we found no significant association between visceral adipose tissue macrophage content and systemic markers of inflammation (specifically with ESR, CRP, Troponin, D-dimer, IL-6, and Ferritin). To our knowledge, this is the first study to directly examine visceral adipose tissue and systemic inflammation among patients who died of COVID-19. Adiposity is an established risk factor for the hyperinflammatory syndrome associated with COVID-19, especially among younger patients, and further studies are needed to elucidate what mechanisms may drive this. However, studies looking for the origins of systemic inflammation may have to look elsewhere than visceral adipose tissue macrophages.

## Data Availability

All data produced in the present study are available upon reasonabl erequest to the authors

